# Physicians’ Reactions to COVID-19: The Results of an International Internet Survey

**DOI:** 10.1101/2021.01.13.21249460

**Authors:** Parul Aneja, Inderjit Singh, Bhupinder Singh, Pardeep Singh Kundi, Inderbir Singh, Sanjana Kathiravan, Shubh Mohan Singh

## Abstract

**Objectives:** Physicians across the world have been disproportionately affected by the COVID-19 pandemic. This study was designed and conducted to assess the emotional, cognitive, and behavioural reactions of physicians to the initial phase of the COVID-19 pandemic.

**Materials and methods:** An online survey questionnaire using the google forms platform was constructed by the authors. The items in the questionnaire were based on clinical experience, relevant literature review and discussion with peers. A list of issues that were deemed as essential components of the experience of the pandemic relevant to physicians was arrived at. Thereafter these issues were operationalized into question form and hosted on the google forms platform. The link to this questionnaire was circulated by the authors among their peer groups in the month of April 2020.

**Results:** We received 295 responses and 3 were unusable. Most of the responses were from India, the United States of America, Australia, Canada and the United Kingdom. About 60% of the respondents identified themselves as frontline and had a decade of clinical experience. Most respondents reported being anxious due to the pandemic and also observed the same in their peers and families. A majority also observed changes in behaviour in self and others and advanced a variety of reasons and concerns. A sense of duty was the most commonly employed coping mechanism.

**Conclusion:** Physicians are not immune from information and misinformation, or cues in the environment. Behavioural choices are not always predicted by knowledge but by a combination of knowledge, emotional state, personality and environment. Healthcare settings need to be ready for emergencies and should focus on reducing uncertainty in physicians. These factors may also be gainfully used in the mental health promotion of physicians in COVID-19 care roles.

## Introduction

The COVID-19 pandemic is a unique worldwide disruptive event. Most people living across almost all countries of the world have been affected by it. Physicians across the world are no exception to the impact of the pandemic. Many have suffered disproportionately with regards to morbidity and mortality and have had to make changes in their work protocols owing to the demands imposed by the pandemic. It is possible that many of these changes are a source of stress in this group of health care workers (HCW).

There are some studies that have assessed mental health consequences and stress in HCW and have shown that there is a possibility of deleterious mental health consequences in this population [1]. Most of these studies are based on HCW as a single bloc, have assessed specific questions such as levels of stress or psychological symptoms and have been conducted in specific geographical areas [2,3].

We believe that the perceived experiential aspects of the pandemic are important determinants in understanding the social, economic and the health consequences of the same. Perceived experience may also impact the reactions to the pandemic. It is also likely that these experiential aspects differ across classes of HCW. However, the experiential effects of the COVID-19 pandemic are not studied as well. We decided to conduct this study to assess the physician perceived experiential effects of living and working through the COVID-19 pandemic. We chose physicians for this study because they are usually in leadership positions in planning and in direct healthcare. Another important reason also is that physicians are possibly at the frontline with regards to possible deleterious consequences of the pandemic with regards to health, social and economic fallouts. Finally the international nature of the proposed survey would enable us to compare the experience in different countries experiencing different trajectories with regards to the effects of the pandemic.

## Material and methods

The study was conducted in the form of an internet survey. This survey was designed to be answered by physicians across the world. We defined physicians as any medical doctor with a completed basic medical degree such as MBBS or MD and working in biomedicine in some capacity. We defined a frontline physician as one who self-identifies as being involved in direct medical care and/or handling biological specimens of known or suspected patients with COVID-19. Years elapsed between the latest medical degree and answering the survey was taken as the years of experience.

Institutional Ethics Committee (intramural) of the PGIMER Chandigarh approval was sought and ethical approval was given for the study (NK/6212/Study/410). The study was designed as an internet survey questionnaire. The items in the questionnaire were devised keeping in mind ease of administration on an internet platform. The investigators used their experience, literature review and discussion with peers to come up with a list of issues that were deemed as essential components of the experience of the pandemic. Thereafter these issues were operationalized into question form. These questions or items could be in the form of multiple choice answers with single or multiple possible responses and also in written paragraphs. These questions went through various rounds of consultation amongst the investigators before we arrived at a consensus survey questionnaire. This was then piloted on 10 volunteer physicians from amongst our peers. Using the feedback, we arrived at the final survey instrument. This instrument was operationalised on the google forms platform. The link along with an invitational message was circulated through text messages and messaging apps through informal and formal channels to likely participants. Each questionnaire took around 10 minutes to complete.

Participation in the study was voluntary. We accepted online responses to the survey from 5th to 23rd April 2020.

## Results

295 responses were received during the study period.

Table 1 presents the sociodemographic profile of the study respondents. 3 responses were incomplete and were excluded. Inferential statistics were done and there were no statistically significant differences in the frequency of different responses across gender, and frontline/non-frontline status.

**Table 1.** Profile of the respondents

| Country | Male | Female | Total (%) | Mean age in years (SD) | Mean years of experience (SD) | Self-identified as frontline |
| --- | --- | --- | --- | --- | --- | --- |
| India | 114 | 60 | 174 (58.82) | 43.51 (8.63) | 15.10 (8.61) | 91 (52.29) |
| USA | 26 | 15 | 41 (14.23) | 44.22 (7.99) | 12.74 (9.14) | 29 (69.04) |
| Canada | 25 | 7 | 32 (10.84) | 47.84 (10.14) | 14.87 (10.49) | 25 (78.12) |
| Australia | 13 | 10 | 23 (7.79) | 43.56 (5.53) | 12.91 (7.37) | 15 (65.21) |
| UK | 11 | 5 | 16 (5.42) | 44.93 (8.85) | 19.56 (9.19) | 13 (81.25) |
| Others | 2 | 4 | 6 (2.03) | 44.16 (6.88) | 16.16 (7.13) | 4 (66.66) |
|  | 191 | 101 | 292 (98.60) | 44.18 (8.54) | 14.85 (8.87) | 177 (60.00) |

A majority of the respondents considered themselves to be well informed about the facts of COVID-19 and this was reflected in the endorsement of correct response in information items in the questionnaire (∼80-94% correct responses). The most common source of information about COVID-19 was from official validated websites such as those of WHO, CDC, NHS etc (91.12% of respondents) followed by social media (39.96%). However a majority of respondents identified multiple sources of information of both formal and informal nature. Only 25.42% respondents identified a single source of information (most commonly through websites such as mentioned above).

We asked the respondents to write down the first word/phrase that came to their mind when they thought of COVID-19. We generated a word cloud from an online resource. The result is presented in figure 1.

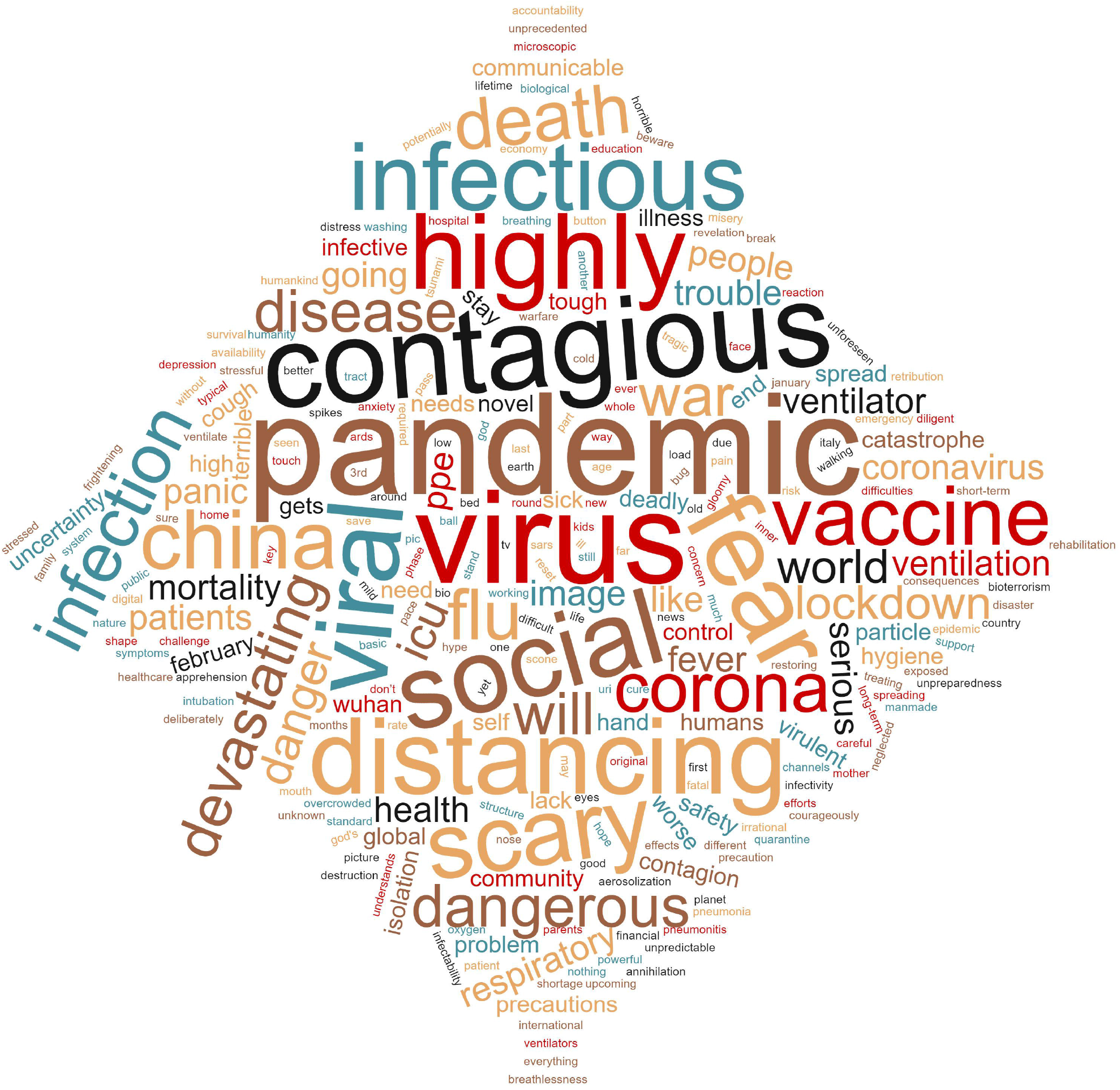

The predominant emotional state as perceived by the respondents is presented in table 2. In addition 61.45% respondents considered the emotional state of people around them to be ‘contagious’ in that it influenced others as well. 27% of the respondents thought that it may be possible. 4.8% of the respondents thought that there was no fear or panic with regards to COVID-19. 87.70% of the respondents thought that there was widespread panic in the society with regards to COVID-19. Only 9.20% of respondents thought that was no panic and fear among health care providers in their areas.

**Table 2.**
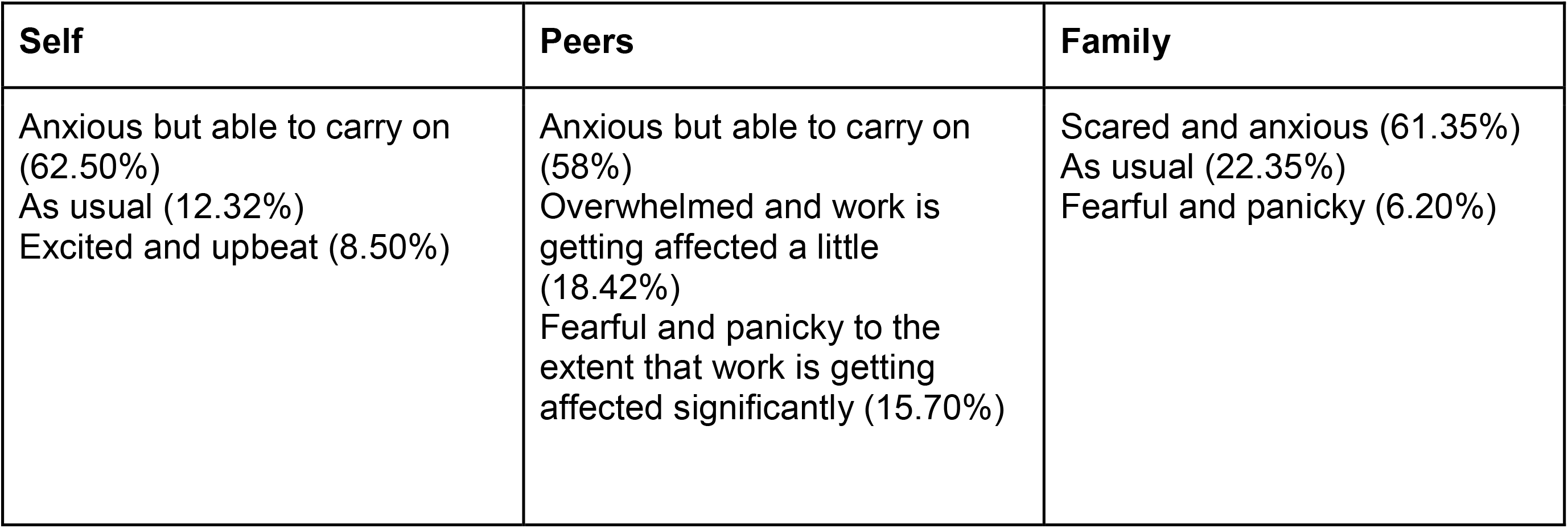
Perceived emotional state of self, peers and family during the study period (top 3 choices endorsed)

| **Self** | **Peers** | **Family** |
| --- | --- | --- |
| Anxious but able to carry on (62.50%)<br>As usual (12.32%)<br>Excited and upbeat (8.50%) | Anxious but able to carry on (58%)<br>Overwhelmed and work is getting affected a little (18.42%)<br>Fearful and panicky to the extent that work is getting affected significantly (15.70%) | Scared and anxious (61.35%)<br>As usual (22.35%)<br>Fearful and panicky (6.20%) |

The changes in behaviour observed by the respondents in self and peers is presented in table 3. Most respondents endorsed more than one change in behaviour and only 8.13% of the respondents did not observe any change. Excessive consumption of and forwarding of COVID-19 related material was the most common change in behaviour noted. Most respondents also had more than a single reason for the changes. The most common reason for the change in behaviour was risk aversion (53.45%) followed by perceived scientific evidence for changes in behaviour (49.70%). Some were influenced by what others had told them (22.90%) or what they had observed in others (18.50%).

**Table 3.** Most commonly observed behavioural changes in self and others (in order of frequency of endorsement)

|  |  |
| --- | --- |
| Sharing and forwarding COVID-19 related information on social media | 67.23% |
| Practicing excessive safety measures while conducting procedures on patients unlikely to be infected unlike before | 48.13% |
| Excessive shopping for essential household items including grocery and food | 42.03% |
| Trying to avoid even those patients who are unlikely to be infected | 36.94% |
| Avoiding patients likely to be infected | 36.90% |
| Hoarding masks, PPEs and/or hand sanitizers | 28.20% |
| Self-medication with medicines such as hydroxychloroquine when not indicated | 19.32% |
| Repeatedly checking for symptoms of COVID-19 in self and family members | 17.96% |
| No change | 8.13% |

The emotional, psychological and physiological changes noticed by the respondents are presented in table 4. More than half of the respondents noticed a variety of changes. The most common was anxiety related to and avoidance of COVID-19 related workplaces.

**Table 4.**
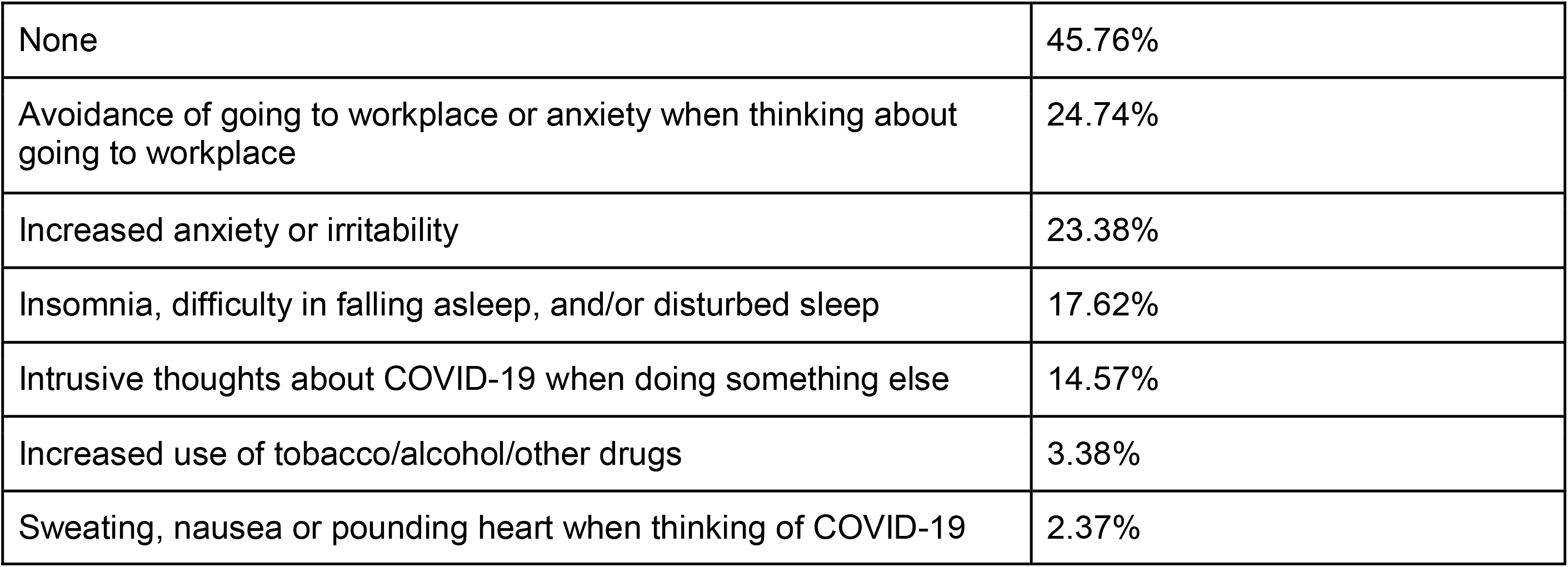
Emotional, psychological and physiological changes in respondents (in order of frequency of endorsement)

|  |  |
| --- | --- |
| None | 45.76% |
| Avoidance of going to workplace or anxiety when thinking about going to workplace | 24.74% |
| Increased anxiety or irritability | 23.38% |
| Insomnia, difficulty in falling asleep, and/or disturbed sleep | 17.62% |
| Intrusive thoughts about COVID-19 when doing something else | 14.57% |
| Increased use of tobacco/alcohol/other drugs | 3.38% |
| Sweating, nausea or pounding heart when thinking of COVID-19 | 2.37% |

The most common fear endorsed by the respondents was to pass on the infection to family members (66.80%), possibility of isolation and quarantine (22.30%), loss of income (19.25%), and death (18.8%). 17.50% of the respondents did not have any concerns and were glad to be of use to the society.

The positive motivational factors as endorsed by the respondents is presented in table 5. In addition, 2 respondents also mentioned their faith in god as their biggest source of motivation. The real-life problems encountered by the respondents are presented in table 6. The most common problems were the lack of personal protective equipment and lack of clarity in administrative setups. There was also a craving for interrupted personal relationships. The experience of stigma was relatively uncommon.

**Table 5.** Positive motivational aspects of working through the COVID-19 pandemic (in order of frequency of endorsement)

|  |  |
| --- | --- |
| Sense of duty and work (it's my job and I am doing it) | 41.69% |
| Intellectual (humanity has been living with all kinds of microorganisms throughout evolution, infection is probably inevitable) | 28.47% |
| Validation of training and experience (It's a test of all my years of training and experience) | 10.84% |
| Novelty (This is the event of our lifetimes and I am not going to sit this one out) | 10.16% |
| Social validation (My family/community/peers are proud of me, can't let them down) | 3.05% |
| None of the above | 3.10% |

**Table 6.** Real life difficulties experienced by respondents (in order of frequency of endorsement)

|  |  |
| --- | --- |
| Non- availability of personal protective equipment, other essential supplies | 69.50% |
| Confusion in workplace administrative setups | 44.40% |
| Less interaction with friends due to social distancing | 41.01% |
| Confined indoors | 40.33% |
| Unable to meet family and friends | 40.33% |
| Stigma from colleagues/community and family/friends for being a potential carrier of infection | 9.49% |
| Too much work with no respite | 6.77% |
| No problems | 5.76% |
| Strained relationships with spouse, family or friends | 4.40% |

The respondents expressed that they could make better behavioural choices for themselves and the community if there was a reduction of uncertainty with regard to availability of protective equipment, food, security, etc. (71.20%), better knowledge (64.40%), more responsive administrative setups (57.28%) and if there were positive role models (49.15%).

## Discussion

The COVID-19 pandemic is a unique event that has not been experienced by humans alive today. It has necessitated widespread changes in the practice of medicine all over the world. It is also increasingly obvious that in the absence of a vaccine or treatment, this pandemic will have a prolonged course in different parts of the world. Thus many of these changes are likely to be long lasting and would require significant adaptations [4]. There may also be different factors involved in coping and motivation in health care providers [5].

This survey reflects the experience of physicians around the world in the initial part of the pandemic. We chose to include only physicians in this sample as they are often in decision making and responsible leadership positions in their practices. Hence their experience of the pandemic is likely to be different from other categories of health care providers [6]. We wanted to assess the experience of physicians across the world and hence examine commonalities and differences if any. We chose to conduct a quantitative study rather than a qualitative one to get a broader impression of the experiences as we also did not want to confine ourselves to any one specific aspect of the experience. For these reasons, an anonymous online survey using a pragmatically derived questionnaire was considered the most effective method of reaching out to a large number of respondents [7]. At the time that this survey was conducted, countries in Europe and the United States were already significantly affected in form of morbidity and mortality. Countries such as India had low prevalence but were already experiencing psychosocial consequences due to the lockdown and misinformation and physicians were involved in preparation for the oncoming pandemic. Hence all countries were experiencing different forms of consequences of the COVID-19 pandemic.

Our sample consisted of physicians in various frontline and non-frontline roles from all over the world. Around 2/3rd of the respondents were male and 60% of the respondents identified themselves as frontline. The mean years of experience suggests that most were in mid-level clinical positions. They were knowledgeable about the facts of COVID-19. Thus we were able to get a fairly representative sample but with a preponderance of respondents from India. However, we did not find any statistically meaningful differences between males and females and frontline/non-frontline doctors. Hence we are discussing the results for the whole population. The world cloud represents the predominant concern as expressed in a single word or phrase with relation to the situation. The most frequently used words were pandemic, virus, dangerous, scary, contagious, China, distancing among others. The main concerns were about the nature and etiology of the pandemic, and the perceived dangerousness and behavioural measures required. The word pandemic seemed to have a special significance as it is a novel emotional and cognitive experience for all living and working through it.

We believe that the prevailing emotional state, behavioural and cognitive changes in the setting of COVID-19 are associated with each other. While anxiety and depressive symptoms have been reported in health care providers in general, we found that this was common in physicians as well [8]. Physicians were receptive to multiple sources of information which include social media and other legitimate websites. It is likely that they may have been exposed to unvalidated information on social media as well that may have influenced their attitudes and behaviours as seen in other populations as well [9]. The use and influence of social media in this population deserves special attention. This is because in spite of better knowledge, physicians observed in self and peers various unexpected behaviours such as mentioned in table 3. A similar dissociation between knowledge and behaviour has been reported in the past as well [10,11]. Similar findings were seen in midwives [12]. In addition, physicians were able to perceive anxiety and negative affect in peers and family to the extent of it being disabling and may have found it contagious. The common reasons advanced for these behaviours were also associated with risk aversion and due to observation of others. A further evidence of the changed emotional state and heightened tension in view of the prevailing situation is presented in table 4 which shows a high prevalence of avoidance, hyperarousal, and intrusive thoughts. Similar effects have been observed in health care providers facing a pandemic elsewhere [13]. Behavioral choices in pandemics are an outcome of knowledge and prevailing emotional state which in turn is influenced by the cues in the environment and cues of anxiety, worry and hyperarousal in self, and personality attributes such as suggestibility and harm avoidance.

The motivational aspects mostly mirrored aspects that have been described earlier [5]. Different aspects may play a differential role in individuals. In our study, sense of duty and intellectual reasons were found to be important factors. The common problems perceived by the physicians were the non-availability of personal protective equipment (PPE), the inability of systems to cope with the novel situation. While the rational use of PPE especially in the initial stages when there were widespread shortages was debated, there is no doubt that PPE was a major concern around the world in all classes of healthcare providers [14]. The problems at the workplace were also often reported [5]. The pandemic caught healthcare systems everywhere off guard and led to the need of rapid adaptation. COVID-19 underlines the need to be ready for major eventualities and also the major impact that uncertainty with regards to personal safety has on morale in healthcare providers in general and physicians in particular. This was reflected in the suggestions given by the respondents with regards to how they could make better behavioural choices.

Our study was able to delineate various important factors that have a bearing of morale of physicians in a pandemic situation. In today’s hyper-connected world, physicians are not immune from information and misinformation, or cues in the environment. Behavioural choices are not always predicted by knowledge but by a combination of knowledge, emotional state, personality and environment. Healthcare settings need to be ready for emergencies and should focus on reducing uncertainty in physicians. These factors may also be gainfully used in the mental health promotion of physicians in COVID-19 care roles.

## Data Availability

Data not available

## Acknowledgement (if applicable)

None of the authors have any acknowledgments to make Funding/financial disclosure (if applicable): None, this was a non-funded study

Competing interests (if applicable): None of the authors have any competing interests to declare

## References

[1] Lima CKT, Carvalho PM de M, Lima I de AAS, Nunes JVA de O, Saraiva JS, de Souza RI, et al. The emotional impact of Coronavirus 2019-nCoV (new Coronavirus disease). Psychiatry Res 2020;287:112915. https://doi.org/10.1016/j.psychres.2020.112915.

[2] Neto MLR, Almeida HG, Esmeraldo JD, Nobre CB, Pinheiro WR, de Oliveira CRT, et al. When health professionals look death in the eye: the mental health of professionals who deal daily with the 2019 coronavirus outbreak. Psychiatry Res 2020;288:112972. https://doi.org/10.1016/j.psychres.2020.112972.

[3] Rajkumar RP. COVID-19 and mental health: A review of the existing literature. Asian J Psychiatry 2020;52:102066. https://doi.org/10.1016/j.ajp.2020.102066.

[4] Barnett DJ, Rosenblum AJ, Strauss-Riggs K, Kirsch TD. Readying for a Post-COVID-19 World: The Case for Concurrent Pandemic Disaster Response and Recovery Efforts in Public Health. J Public Health Manag Pract JPHMP 2020;26:310–3. https://doi.org/10.1097/PHH.0000000000001199.

[5] Mohindra R, R R, Suri V, Bhalla A, Singh SM. Issues relevant to mental health promotion in frontline health care providers managing quarantined/isolated COVID19 patients. Asian J Psychiatry 2020;51:102084. https://doi.org/10.1016/j.ajp.2020.102084.

[6] Zhang L, Li B, Jia P, Pu J, Bai B, Li Y, et al. [An analysis of global research on SARS-CoV-2]. Sheng Wu Yi Xue Gong Cheng Xue Za Zhi J Biomed Eng Shengwu Yixue Gongchengxue Zazhi 2020;37:236–45. https://doi.org/10.7507/1001-5515.202002034.

[7] Geldsetzer P. Use of Rapid Online Surveys to Assess People’s Perceptions During Infectious Disease Outbreaks: A Cross-sectional Survey on COVID-19. J Med Internet Res 2020;22:e18790. https://doi.org/10.2196/18790.

[8] Lai J, Ma S, Wang Y, Cai Z, Hu J, Wei N, et al. Factors Associated With Mental Health Outcomes Among Health Care Workers Exposed to Coronavirus Disease 2019. JAMA Netw Open 2020;3:e203976. https://doi.org/10.1001/jamanetworkopen.2020.3976.

[9] González-Padilla DA, Tortolero-Blanco L. Social media influence in the COVID-19 Pandemic. Int Braz J Urol Off J Braz Soc Urol 2020;46. https://doi.org/10.1590/S1677-5538.IBJU.2020.S121.

[10] Garfin DR, Silver RC, Holman EA. The novel coronavirus (COVID-2019) outbreak: Amplification of public health consequences by media exposure. Health Psychol Off J Div Health Psychol Am Psychol Assoc 2020;39:355–7. https://doi.org/10.1037/hea0000875.

[11] Corley A, Hammond NE, Fraser JF. The experiences of health care workers employed in an Australian intensive care unit during the H1N1 Influenza pandemic of 2009: a phenomenological study. Int J Nurs Stud 2010;47:577–85. https://doi.org/10.1016/j.ijnurstu.2009.11.015.

[12] Sögüt S, Dolu İ, Cangöl E. The relationship between COVID-19 knowledge levels and anxiety states of midwifery students during the outbreak: A cross-sectional web-based survey. Perspect Psychiatr Care 2020. https://doi.org/10.1111/ppc.12555.

[13] Wang C, Pan R, Wan X, Tan Y, Xu L, Ho CS, et al. Immediate Psychological Responses and Associated Factors during the Initial Stage of the 2019 Coronavirus Disease (COVID- 19) Epidemic among the General Population in China. Int J Environ Res Public Health 2020;17. https://doi.org/10.3390/ijerph17051729.

[14] Amerio A, Bianchi D, Santi F, Costantini L, Odone A, Signorelli C, et al. Covid-19 pandemic impact on mental health: a web-based cross-sectional survey on a sample of Italian general practitioners. Acta Bio-Medica Atenei Parm 2020;91:83–8. https://doi.org/10.23750/abm.v91i2.9619.

